# The Width–Delay Index: a Glucose-Only OGTT Metric for Assessing Insulin Resistance

**DOI:** 10.64898/2026.07.05.26357315

**Authors:** Ren Zhang

## Abstract

**Background:** Insulin resistance is a core pathophysiologic feature of metabolic disease, but its reference-standard assessment by steady-state plasma glucose (SSPG) testing is procedurally demanding and labor-intensive, limiting use in routine clinical care and large-scale research. Because OGTT glucose profiles are widely available, we aimed to develop a glucose-only metric to characterize dynamic glucose responses and estimate SSPG-measured insulin resistance.

**Methods:** We developed the Width–Delay Index (WDI), a glucose-only OGTT metric integrating relative exposure width, delayed exposure timing, and glycemic floor. In a dataset of 32 subjects with 16-point venous OGTT profiles and paired SSPG measurements, WDI performance was assessed using leave-one-out cross-validation (LOOCV) for SSPG prediction, together with insulin-resistance discrimination and sparse-sampling robustness analyses.

**Results:** The 15–120 min OGTT window yielded the strongest WDI performance. WDI15–120 predicted SSPG with LOOCV *R^2^* =0.57 (95% CI, 0.27–0.77), Pearson *r* = 0.77, and Spearman *ρ* = 0.74. WDI15–120 outperformed standard OGTT glucose measures and insulin-derived indices, including HOMA-IR, Matsuda index, and disposition index. WDI15–120 also discriminated insulin-resistant from insulin-sensitive subjects with AUROC = 0.969. When recalculated from conventional 5-point OGTT sampling, WDI15–120 retained substantial performance, with LOOCV *R^2^* = 0.41 and AUROC = 0.945.

**Conclusions:** WDI provides a simple, glucose-only, physiologically interpretable approach for estimating SSPG-measured insulin resistance from OGTT glucose dynamics.

## Introduction

Insulin resistance is a central metabolic abnormality underlying type 2 diabetes, obesity-related metabolic dysfunction, and cardiometabolic disease risk (Accili, et al., 2025; Li, et al., 2022; Petersen and Shulman, 2018). Accurate assessment of insulin resistance is important for metabolic phenotyping, risk stratification, and evaluation of metabolic interventions (Khamseh, et al., 2024; Piché, et al., 2020). Controlled physiological methods provide experimental assessment of insulin-mediated glucose disposal (Gastaldelli, 2022). One such method is the insulin suppression test, in which endogenous insulin secretion is suppressed while insulin and glucose are infused to create comparable steady-state insulin exposure across individuals.

Under these conditions, the resulting steady-state plasma glucose (SSPG) concentration reflects the ability to dispose of glucose in response to insulin: higher SSPG indicates poorer insulin-mediated glucose disposal and greater insulin resistance (Knowles, et al., 2013; Shen, et al., 1970). The euglycemic–hyperinsulinemic clamp provides another established approach, using glucose infusion requirements during controlled hyperinsulinemia to estimate insulin-stimulated glucose disposal (DeFronzo, et al., 1979; Gastaldelli, 2022). Although these methods provide direct and accurate physiological assessment, they are labor-intensive, time-consuming, and difficult to apply in routine clinical practice or large research cohorts. This has motivated the widespread use of simpler surrogate indices derived from fasting blood tests or oral glucose tolerance test (OGTT) measurements.

These surrogate indices range from simple glucose measurements to insulin-based composite indices. Standard OGTT interpretation commonly uses fasting glucose, 1-h glucose, 2-h glucose, peak glucose, mean glucose, or total glucose AUC. These measures are clinically accessible, but they summarize the glucose response as isolated concentrations or aggregate exposure and do not explicitly quantify whether post-load glucose exposure is narrow and rapidly resolving or broad and delayed. Insulin-derived indices provide additional physiological information: HOMA-IR reflects fasting glucose–insulin homeostasis (Matthews, et al., 1985; Tahapary, et al., 2022), whereas the Matsuda index (Matsuda and DeFronzo, 1999) and disposition index (Kahn, et al., 1993; Utzschneider, et al., 2009) incorporate glucose and insulin responses during the OGTT. However, these indices require insulin measurements, which are not always collected in standard OGTT protocols and may vary across assays and study settings. Although dynamic and model-based OGTT approaches have been developed, many require insulin measurements, curve fitting, or multiple estimated parameters (Metwally, et al., 2026; Metwally, et al., 2025; Tsai, et al., 2024). These limitations highlight the need for a simple, glucose-only, physiologically interpretable OGTT metric that captures dynamic curve physiology, avoids insulin measurements and complex model fitting, and can be directly benchmarked against SSPG.

The OGTT glucose curve contains information beyond isolated glucose values or total exposure. In particular, impaired insulin-mediated disposal would be expected to prolong post-load glucose exposure and shift more of that exposure later in the test. We therefore developed the Width–Delay Index (WDI) as a simple, glucose-only metric designed to capture broad and delayed glucose exposure in a physiologically interpretable form. Using a well-characterized dataset with dense 16-point venous OGTT profiles paired with SSPG measurements, we tested whether WDI could identify an informative OGTT analysis window, predict SSPG-measured insulin resistance from glucose values alone, compare favorably with conventional glucose and insulin-derived indices, discriminate insulin-resistant from insulin-sensitive subjects, and remain robust under conventional sparse OGTT sampling.

## Materials and Methods

### Dataset and OGTT preprocessing

The analysis used a dense venous OGTT dataset with paired steady-state plasma glucose (SSPG) measurements as the reference measure of insulin resistance (Metwally, et al., 2025). The primary analytical cohort included 32 subjects with SSPG values and sufficient venous glucose measurements to calculate WDI over the primary 15–120 min window. Insulin-sensitive and insulin-resistant group labels were taken from the original SSPG-based classification provided with the dataset. Venous glucose profiles were organized by subject and timepoint.

Duplicate values at the same subject-timepoint were averaged. Glucose values at analysis-window boundaries were taken directly when available or estimated by linear interpolation when bracketed by observed measurements; no extrapolation was performed. Subjects without sufficient coverage for a given window were excluded from that analysis only. This study was a secondary analysis of de-identified human OGTT and SSPG data from a previously published study. Ethical approval and informed consent for the original cohort were obtained as described in the source publication. No new participants were recruited, and no identifiable human data or biospecimens were accessed.

### Width–Delay Index definition and calculation

WDI was calculated for a specified OGTT analysis window *W* = [*s*, *e*], where *s*and *e*denote the start and end times of the window. Fasting glucose was defined as *G*_0_ = *G*(0). Within each window, *G*_max_was defined as the maximum glucose value and *G*_low_as the minimum glucose value. Glucose exposure above fasting was calculated as:

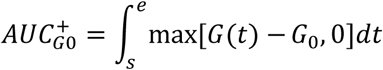

using trapezoidal integration. The relative width term was calculated as:

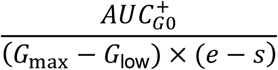

This term represents a dimensionless estimate of the temporal breadth of the glucose excursion after normalization to the curve’s peak-to-floor amplitude. To quantify delayed exposure, glucose was first expressed relative to the within-window glycemic floor:

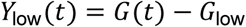

Cumulative exposure above *G*_low_was then calculated over the window. *T*50_low_was defined as the time at which cumulative exposure reached 50% of total exposure above *G*_low_. The normalized delay term was:

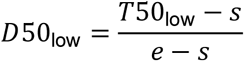

Finally, WDI was calculated as:

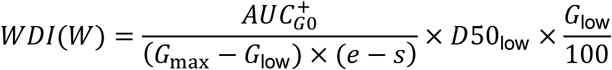

Thus, WDI combines relative glucose-exposure width, delayed exposure timing, and glycemic floor scaling. For the primary analysis, WDI was calculated over the 15–120 min window and denoted WDI15–120.

### SSPG prediction, window selection, and comparator metrics

SSPG was used as the reference measure of insulin resistance. For each WDI window or comparator metric, SSPG was predicted using univariate linear regression with the metric as the sole predictor. Performance was evaluated by leave-one-out cross-validation (LOOCV), in which each subject was predicted once after fitting the model in all remaining subjects. LOOCV *R*^2^was the primary performance metric, and RMSE quantified prediction error in SSPG units. For window selection, the same WDI formula was applied across candidate OGTT windows, and the window with the highest LOOCV *R*^2^was selected for subsequent analyses. WDI15–120 was then compared with standard glucose metrics, including fasting, 30-min, 1-h, 90-min, 2-h, peak, mean glucose, and total AUC, and with insulin-derived indices including HOMA-IR, Matsuda index, and disposition index. Pearson and Spearman correlations were calculated in the native direction of each metric.

### Insulin-resistance classification analysis

Insulin-sensitive and insulin-resistant groups were defined using the SSPG-based labels provided with the original dataset. WDI15–120 values were compared between groups using a two-sided Mann–Whitney test and summarized as median [IQR]. Binary discrimination was evaluated by receiver operating characteristic analysis, with AUROC used as the primary classification metric. For metrics inversely related to insulin resistance, such as Matsuda index, the ROC direction was reversed so that higher classification scores corresponded to greater insulin resistance.

### Sparse-sampling robustness analysis

To assess whether WDI15–120 could be estimated from reduced OGTT sampling, we recalculated WDI using selected sparse schedules, including 0/30/60/90/120, 0/30/60/120, and 0/30/90/120 min. The WDI formula was kept fixed as WDI15–120 for all schedules. For each sparse schedule, only the retained glucose samples were used; omitted dense timepoints were not used to calculate *G*_max_, *G*_low_, AUC, *D*50_low_, or WDI. Sparse profiles were treated as piecewise-linear curves connecting the retained samples. When 15 min was not directly sampled, the 15-min boundary value was estimated by linear interpolation from adjacent retained samples. All WDI components were then recalculated from the sparse 15–120 min curve and evaluated for SSPG prediction using the same LOOCV framework as the dense analysis.

### Statistical analysis and software

Analyses were performed in Python using numpy, pandas, scipy, and scikit-learn. Continuous prediction of SSPG was evaluated using univariate linear regression with LOOCV. Confidence intervals for LOOCV *R²* and RMSE were estimated using nonparametric subject-level bootstrap resampling. For each metric, paired observed SSPG values and leave-one-out cross-validated predicted SSPG values were resampled with replacement 10,000 times. LOOCV *R²* and RMSE were recalculated in each bootstrap resample, and 95% confidence intervals were defined as the 2.5th and 97.5th percentiles of the bootstrap distribution. Model performance was reported as LOOCV *R*^2^ and RMSE. Pearson correlation was used to assess linear association with SSPG, and Spearman correlation was used to assess rank-based association. Group differences between insulin-sensitive and insulin-resistant subjects were tested using the two-sided Mann–Whitney test and summarized as median [IQR]. Binary discrimination was assessed using AUROC. Statistical tests were two-sided, and *p* < 0.05 was considered statistically significant. Figures were generated using Python/matplotlib.

## Results

### Width–Delay Index conceptual framework and construction

During an OGTT, the plasma glucose profile reflects the balance between glucose appearance from intestinal absorption and glucose removal through insulin-mediated disposal. After oral glucose ingestion, absorbed glucose enters the circulation over time, while insulin secretion promotes glucose uptake, particularly into skeletal muscle, and suppresses endogenous glucose production. In insulin-sensitive individuals, this disposal response efficiently offsets glucose appearance, so post-load glucose exposure is relatively brief and resolves earlier. In insulin-resistant individuals, impaired insulin-mediated disposal slows the clearance phase. As a result, glucose exposure persists across a larger fraction of the OGTT period, producing a broader excursion, and a greater portion of the exposure occurs later in the test, producing delayed exposure timing (Fig. 1A). This physiological pattern motivated WDI as a glucose-only index designed to quantify both the width and delay of post-load glucose exposure.

**Fig. 1.**
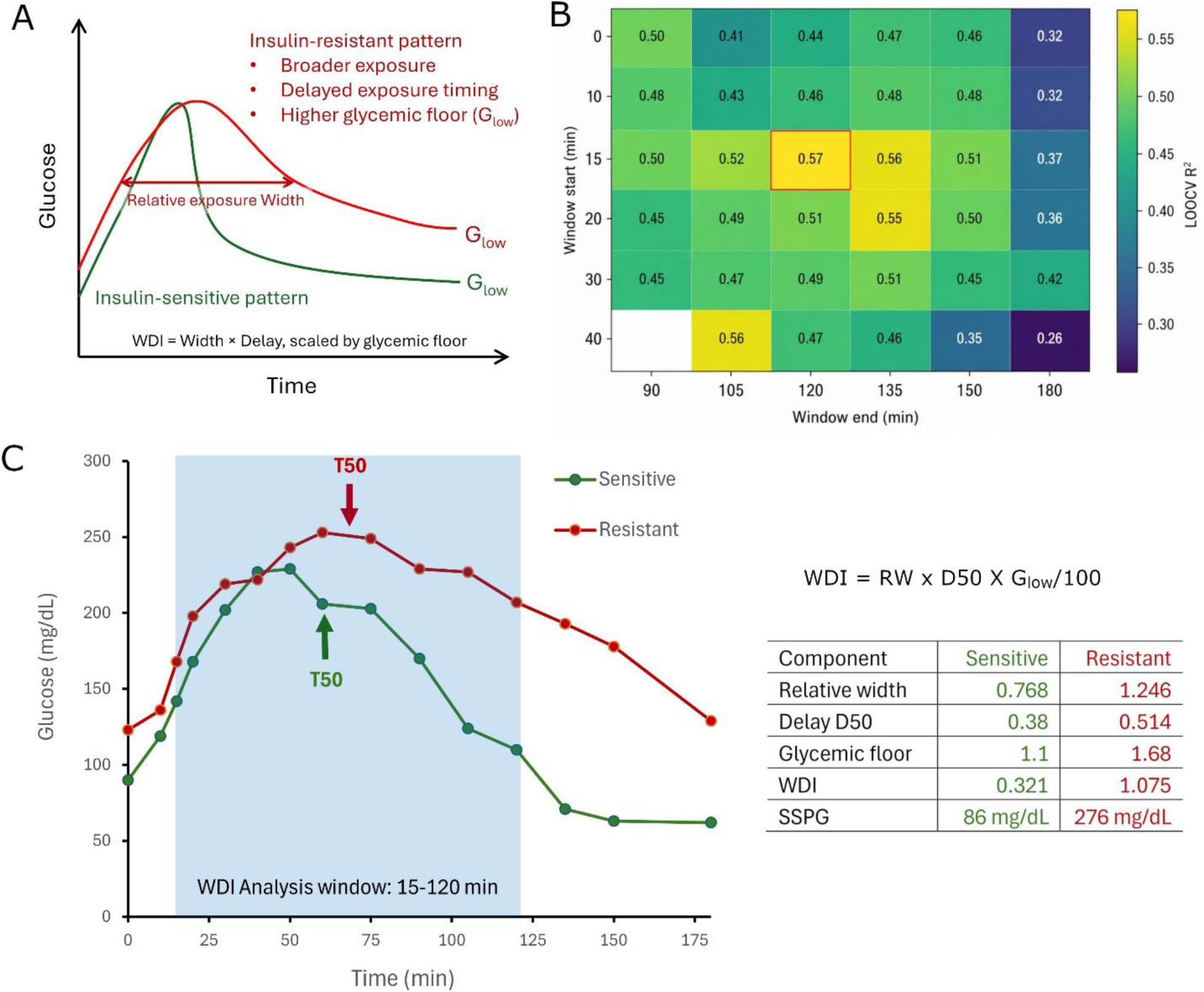
Conceptual framework of the Width–Delay Index and its component decomposition. A) Schematic representation of oral glucose challenge profiles associated with insulin sensitivity and insulin resistance. The insulin-resistant profile shows broader glucose exposure, delayed exposure timing, and a higher glycemic floor. The glycemic floor (G_low_) is the lowest glucose level within the analysis window. WDI integrates relative exposure width and delay, scaled by the glycemic floor. B) Window analysis showing leave-one-out cross-validated *R*^2^values for WDI across OGTT analysis windows. The 15–120 min window showed the strongest performance. (C) Representative OGTT glucose profiles from an insulin-sensitive subject (S42, SSPG = 86 mg/dL) and an insulin-resistant subject (S38, SSPG = 276 mg/dL) within the 15–120 min WDI analysis window. Arrows indicate *T*50*_low_*, the time at which 50% of glucose exposure above the within-window glycemic floor has accumulated. *D*50*_low_*is the normalized timing used in WDI, calculated as (*T*50*_low_* − *s*)/(*e* − *s*). The component table decomposes WDI into relative width, *D*50*_low_*, and glycemic scaling (*G_low_*/100), illustrating higher WDI in the insulin-resistant profile.

WDI quantifies this pattern using three components. The first component is a relative width term, intended as a simple, parameter-light estimate of how broad the post-load glucose excursion is. Rather than fitting a curve shape or estimating multiple kinetic parameters, WDI uses the geometric relationship that AUC reflects both amplitude and temporal spread. This term is calculated as 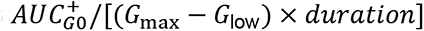, where 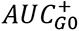 is glucose exposure above fasting. For a given excursion amplitude, a narrow spike contributes relatively little AUC, whereas a broad or plateau-like excursion contributes more AUC because exposure is distributed across a larger portion of the test. Therefore, dividing 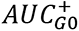 by the peak-to-floor glucose range, *G*_max_ − *G*_low_, reduces the amplitude contribution and yields a width-like measure of temporal spread. Dividing again by test duration converts this into a dimensionless relative width. Thus, this term does not represent the actual duration of hyperglycemia; it provides a normalized estimate of whether the glucose excursion is narrow and transient or broad and persistent.

The second component measures delayed glucose exposure using *D*50_low_. To calculate this term, glucose exposure is first expressed relative to the glycemic floor, *G*(*t*) − *G*_low_. *T*50_low_is then defined as the time at which cumulative exposure above *G*_low_reaches 50% of the total exposure, and *D*50_low_is calculated as (*T*50_low_ − *s*)/(*e* − *s*), where *s* and *e* are the start and end of the analysis period. Thus, *D*50_low_ represents the normalized temporal midpoint of glucose exposure above the glycemic floor. When glucose exposure occurs early and resolves quickly, *D*50_low_ is lower; when exposure persists later in the OGTT, *D*50_low_ increases. Therefore, this term captures delayed exposure timing rather than simply the time of peak glucose.

The third component is a glycemic floor scaling term, *G*_low_/100. While the relative width and *D*50_low_ terms describe the shape and timing of glucose exposure, they are largely normalized features. Two OGTT profiles could therefore have similar width-delay patterns but differ in the absolute glycemic level at which that pattern occurs. *G*_low_, defined as the lowest glucose value within the analysis period, captures the lower envelope of the glucose curve. A higher *G*_low_ indicates that glucose remains elevated even at the lowest point of the profile, consistent with a more persistently elevated glycemic state. Multiplying by *G*_low_/100 therefore scales the width-delay pattern by its glycemic level, so WDI is highest when glucose exposure is not only broad and delayed, but also maintained at a higher floor.

Thus, WDI was defined as the product of relative width, delayed exposure timing, and glycemic floor scaling:

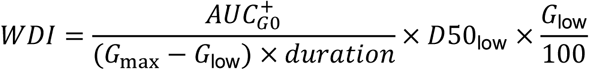

where 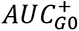 is the glucose AUC above fasting glucose, *G*_max_ and *G*_low_ are the maximum and minimum glucose values within the analysis period, *D*50_low_is the normalized time at which 50% of glucose exposure above *G*_low_ has accumulated, and *duration*is the length of the analysis period. Therefore, WDI increases when post-load glucose exposure is broader, shifted later, and maintained at a higher glycemic floor.

### Window selection and representative profile decomposition

Because OGTT physiology changes over time, we evaluated whether WDI performance depended on the analysis window. Early post-load measurements reflect the transition from fasting to the stimulated state and are influenced by glucose appearance from intestinal absorption and early insulin secretion. Intermediate measurements capture the major period during which insulin-mediated glucose disposal offsets ongoing glucose appearance. Later measurements increasingly reflect recovery-phase physiology, possible glucose undershoot, and additional late-phase variability. Therefore, the most informative interval for WDI may not correspond to the full OGTT duration. We consequently compared WDI across candidate windows to identify the interval that best captured SSPG-related width-delay physiology.

Across candidate windows, WDI performance was evaluated by leave-one-out cross-validation (LOOCV) for SSPG prediction. In this procedure, one subject was held out, the relationship between WDI and SSPG was fit in the remaining subjects, and SSPG was predicted for the held-out subject; this was repeated until every subject had been predicted once. The resulting LOOCV *R*^2^therefore reflects prediction of subjects not used to fit the model, making it a more objective criterion than in-sample correlation for selecting the WDI window. WDI performance varied substantially across windows, indicating that the SSPG-related signal depended on the portion of the OGTT curve analyzed. The 15–120 min window showed the strongest LOOCV *R*^2^and was therefore selected as the primary WDI window for subsequent analyses (Fig. 1B). This interval is also consistent with the intended biology of WDI, because it focuses on the main post-load exposure and clearance period while avoiding the earliest transition phase and later recovery-phase variability.

Using the selected 15–120 min window, representative OGTT profiles illustrated how WDI components differed between insulin-sensitive and insulin-resistant subjects. Compared with the insulin-sensitive subject, the insulin-resistant subject showed a broader glucose excursion, a later *D*50_low_, and a higher *G*_low_, resulting in a higher WDI value (Fig. 1C). This component decomposition illustrates how WDI integrates the temporal spread, timing, and glycemic floor of the post-load glucose profile rather than relying on a single glucose landmark.

### Comparison of WDI15–120 with standard OGTT and insulin-derived indices

After selecting WDI15–120 as the primary index, we compared its performance with standard OGTT glucose metrics and established insulin-derived indices for predicting SSPG. Glucose-only comparators included fasting glucose, 30-min glucose, 1-h glucose, 90-min glucose, 2-h glucose, peak glucose, mean glucose, and total AUC. Insulin-derived comparators included HOMA-IR, Matsuda index, and disposition index. For each metric, predictive performance was evaluated using leave-one-out cross-validated *R*^2^ and RMSE, with RMSE representing the typical prediction error in SSPG units. Pearson and Spearman correlations were also calculated to assess subject-level associations with SSPG.

In this benchmark comparison, WDI15–120 showed the strongest SSPG prediction among the evaluated metrics (Fig. 2A; Table 1). WDI15–120 achieved a LOOCV *R*^2^of 0.57 with an RMSE of 49.6 mg/dL, and was strongly associated with SSPG by both Pearson correlation (*r* = 0.77, *p* < 0.001) and Spearman correlation (*ρ* = 0.74, *p* < 0.001). Among insulin-derived indices, HOMA-IR showed the next highest predictive performance (LOOCV *R*^2^ = 0.33, RMSE = 61.6 mg/dL), followed by disposition index and Matsuda index. Conventional glucose-only OGTT metrics showed lower performance, including mean glucose 0–120 min and total AUC 0–120 min (both LOOCV *R*^2^ = 0.20) and 2-h glucose (LOOCV *R*^2^ = 0.18). Thus, WDI15–120 provided stronger SSPG prediction than both standard OGTT glucose summaries and established insulin-derived surrogate indices.

**Fig. 2.**
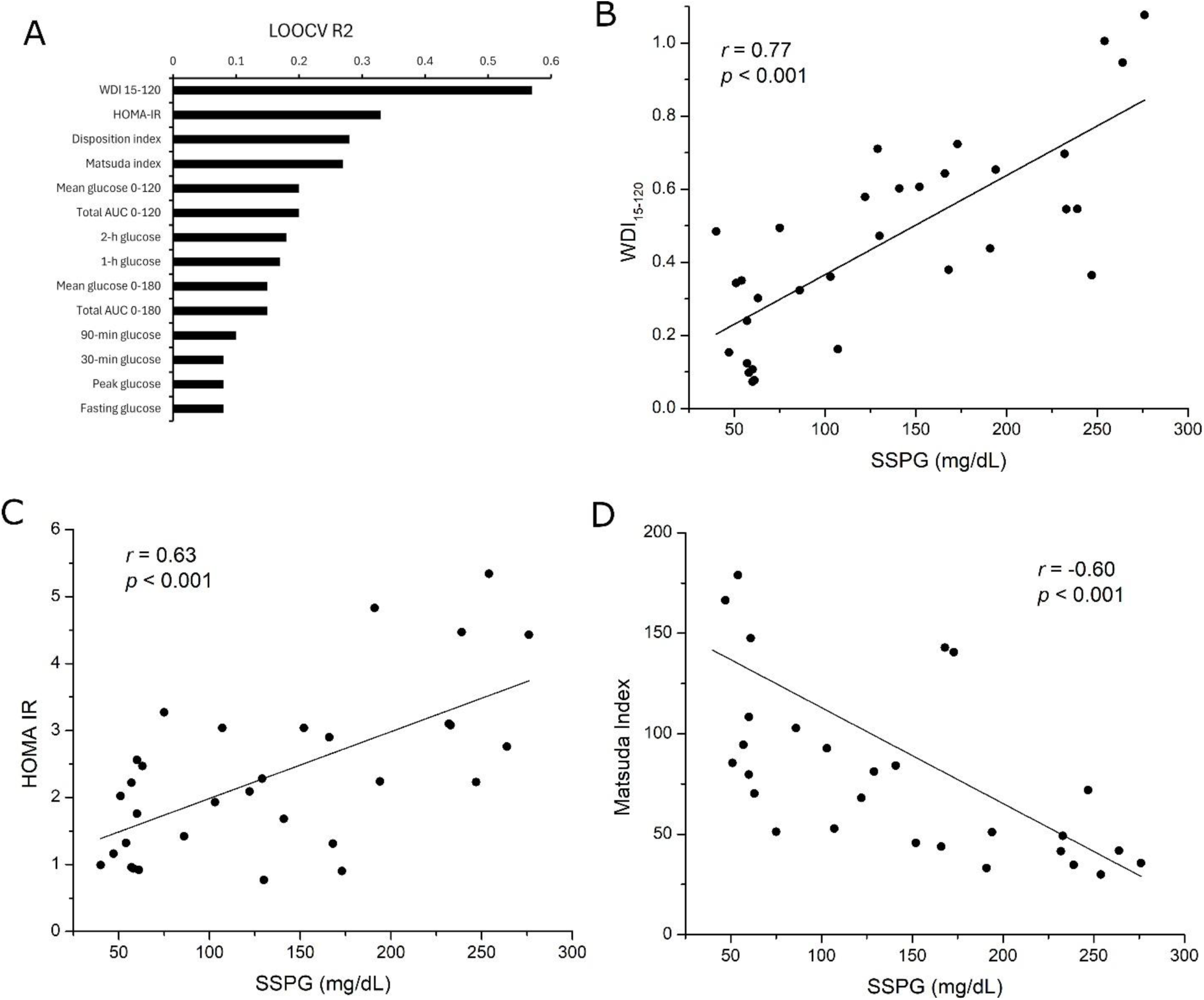
Performance of WDI for predicting SSPG compared with standard OGTT and insulin-derived indices. (A) Leave-one-out cross-validated *R*^2^values for WDI 15–120 and comparator metrics. WDI showed the highest predictive performance for SSPG. (B) Association between SSPG and WDI 15–120. (C) Association between SSPG and HOMA-IR. (D) Association between SSPG and Matsuda index. Pearson correlation coefficients and nominal *p*-values are shown.

**Table 1.** Comparison of WDI_15-120_ with standard OGTT glucose metrics and insulin-derived indices for predicting SSPG.

| Metric | Input | LOOCV $R^2$ | LOOCV $R^2$ (95% CI) | RMSE | RMSE 95% CI | Pearson $r$ | $p$ | Spearman rho | $p$ |
| --- | --- | --- | --- | --- | --- | --- | --- | --- | --- |
| WDI<sub>15-120</sub> | OGTT glucose | 0.57 | 0.27–0.77 | 49.6 | 35.8–62.9 | 0.77 | <0.001 | 0.74 | <0.001 |
| HOMA-IR | Glucose and insulin | 0.33 | –0.03–0.55 | 61.6 | 50.0–72.5 | 0.63 | <0.001 | 0.57 | <0.001 |
| Disposition index | Glucose and insulin | 0.28 | –0.03–0.46 | 64.0 | 51.9–74.9 | –0.60 | <0.001 | –0.56 | <0.001 |
| Matsuda index | Glucose and insulin | 0.27 | –0.15–0.48 | 64.6 | 52.7–75.0 | –0.60 | <0.001 | –0.73 | <0.001 |
| Mean glucose 0-120 | OGTT glucose | 0.20 | –0.21–0.44 | 67.2 | 54.2–79.4 | 0.55 | 0.001 | 0.59 | <0.001 |
| Total AUC 0-120 | OGTT glucose | 0.20 | –0.22–0.45 | 67.2 | 53.9–79.1 | 0.55 | 0.001 | 0.59 | <0.001 |
| 2-h glucose | OGTT glucose | 0.18 | –0.40–0.48 | 68.1 | 52.1–83.3 | 0.57 | <0.001 | 0.62 | <0.001 |
| 1-h glucose | OGTT glucose | 0.17 | –0.20–0.38 | 68.8 | 55.3–81.4 | 0.50 | 0.003 | 0.56 | <0.001 |
| Mean glucose 0-180 | OGTT glucose | 0.15 | –0.37–0.43 | 70.4 | 56.4–84.0 | 0.53 | 0.003 | 0.56 | 0.001 |
| Total AUC 0-180 | OGTT glucose | 0.15 | –0.37–0.43 | 70.4 | 56.3–83.6 | 0.53 | 0.003 | 0.56 | 0.001 |
| 90-min glucose | OGTT glucose | 0.10 | –0.67–0.45 | 71.7 | 53.4–91.2 | 0.54 | 0.001 | 0.64 | <0.001 |
| 30-min glucose | OGTT glucose | 0.08 | –0.33–0.31 | 72.2 | 58.9–84.1 | 0.44 | 0.013 | 0.50 | 0.003 |
| Peak glucose | OGTT glucose | 0.08 | –0.33–0.31 | 72.3 | 59.4–84.3 | 0.45 | 0.010 | 0.53 | 0.002 |
| Fasting glucose | Fasting glucose | 0.08 | –0.30–0.30 | 72.4 | 59.1–84.4 | 0.43 | 0.014 | 0.44 | 0.013 |

Analyses used n=32 subjects unless otherwise indicated. 0–180 min glucose metrics used n=29 because three subjects lacked usable venous glucose coverage through 180 min. LOOCV $R^2$ and RMSE 95% confidence intervals were estimated by subject-level bootstrap resampling using 10,000 resamples. HOMA-IR, Matsuda index, and disposition index were phenotype-table glucose–insulin indices. Correlations are shown in the native metric direction.

The subject-level scatter plots supported these benchmark results. WDI15–120 showed a strong positive relationship with SSPG, consistent with higher WDI values reflecting greater insulin resistance (Fig. 2B). HOMA-IR was also positively associated with SSPG, but with greater dispersion and lower cross-validated predictive performance (Fig. 2C). In contrast, Matsuda index showed the expected inverse association with SSPG, because lower Matsuda values indicate lower insulin sensitivity, but its predictive performance was weaker than WDI15–120 (Fig. 2D). Together, these results indicate that the width-delay pattern captured by WDI15–120 contains SSPG-relevant information beyond conventional glucose summaries and established insulin-derived surrogate indices.

### Discrimination of insulin-resistant and insulin-sensitive subjects by WDI15–120

We next evaluated whether WDI15–120 differed between the insulin-sensitive and insulin-resistant groups defined by the SSPG-based classification used in the original study dataset. WDI15–120 was substantially higher in insulin-resistant subjects than in insulin-sensitive subjects: median 0.60 [IQR 0.53–0.62] versus 0.24 [IQR 0.15–0.38], respectively. The group difference was significant by two-sided Mann–Whitney test (*p* < 0.001; insulin-sensitive *n* = 15, insulin-resistant *n* = 17; Fig. 3A). This separation is consistent with the interpretation that insulin-resistant subjects have broader, later, and higher-floor post-load glucose exposure.

**Fig. 3.**
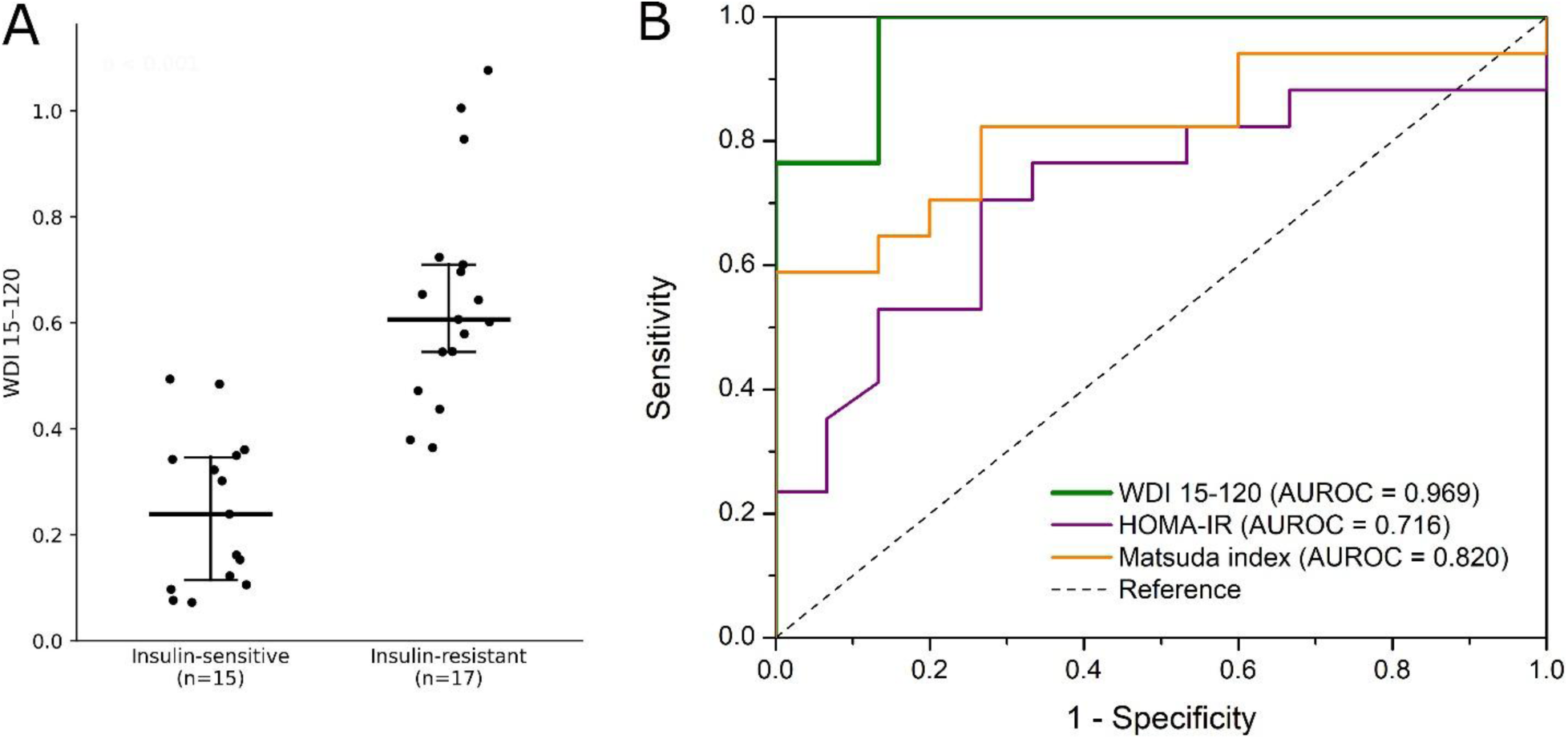
Discrimination of insulin-resistant and insulin-sensitive subjects by WDI. (A) WDI 15–120 values in insulin-sensitive and insulin-resistant subjects. Points represent individual subjects; horizontal bars indicate median and interquartile range. Groups were compared using the Mann–Whitney test. (B) Receiver operating characteristic curves for classification of insulin-resistant versus insulin-sensitive subjects. WDI 15–120 showed stronger discrimination than HOMA-IR and Matsuda index. For Matsuda index, the ROC direction was reversed because lower values indicate greater insulin resistance.

ROC analysis further confirmed strong discrimination of the original insulin-resistance groups by WDI15–120. WDI15–120 achieved an AUROC of 0.969, compared with 0.716 for HOMA-IR and 0.820 for Matsuda index (Fig. 3B). For Matsuda index, the ROC direction was reversed because lower Matsuda values indicate greater insulin resistance. These results indicate that WDI15–120, despite using only OGTT glucose values, discriminated insulin-resistant from insulin-sensitive subjects more strongly than the insulin-derived comparator indices.

### Sparse-sampling robustness of WDI15–120

Although dense OGTT sampling is useful for characterizing glucose dynamics, many clinical and research OGTT protocols use fewer timepoints, commonly including 0, 30, 60, 90, and 120 min or even more limited schedules. We therefore tested whether WDI15–120 could be approximated from reduced OGTT sampling. For each schedule, the WDI formula was kept fixed, and all WDI components were recalculated using only the retained glucose samples. Dense OGTT measurements omitted from a given sparse schedule were not used. Sparse profiles were treated as piecewise-linear curves connecting the retained samples; when 15 min was not directly sampled, the 15-min boundary value was estimated by linear interpolation from adjacent retained samples. Thus, the analysis tested whether the same WDI15–120 construct could be estimated from commonly used sparse OGTT sampling without using information from omitted dense timepoints.

The traditional 5-point OGTT schedule retained meaningful WDI performance. When WDI15–120 was recalculated using only 0, 30, 60, 90, and 120 min glucose values, LOOCV *R²* was 0.41 compared with 0.57 for the dense reference, with RMSE increasing from 49.6 to 57.7 mg/dL (Table 2). Reduced 4-point schedules showed similar performance: the 0/30/60/120 schedule achieved LOOCV *R²* = 0.39, RMSE = 58.9 mg/dL, and AUROC = 0.929, while the 0/30/90/120 schedule achieved LOOCV *R²* = 0.42, RMSE = 57.5 mg/dL, and AUROC = 0.945. These findings indicate that WDI15–120 can be approximated from conventional 5-point OGTT sampling and from selected 4-point schedules with moderate preservation of SSPG-predictive performance. The comparable performance of the two 4-point schedules suggests that reduced sampling can retain meaningful width-delay information when early post-load and late recovery measurements are preserved. Overall, the sparse-sampling analysis supports conventional 5-point OGTT sampling as a feasible reduced-sampling approach for estimating WDI15–120, while indicating that selected 4-point schedules may also provide a practical approximation.

**Table 2.** Sparse-sampling robustness of fixed WDI15–120 for SSPG prediction.

| Sampling analysis | Retained OGTT samples (min) | Curve used for WDI15-120 | LOOCV $R^2$ | RMSE (mg/dL) | Pearson $r$ | Spearman $\rho$ | AUROC |
| --- | --- | --- | --- | --- | --- | --- | --- |
| Dense reference | 0/10/15/20/30/40/50/60/75/90/105/120 | 15/20/30/40/50/60/75/90/105/120 | 0.57 | 49.6 | 0.77 | 0.74 | 0.969 |
| Traditional 5-point OGTT | 0/30/60/90/120 | 15*/30/60/90/120 | 0.41 | 57.7 | 0.69 | 0.67 | 0.945 |
| Reduced 4-point OGTT | 0/30/60/120 | 15*/30/60/120 | 0.39 | 58.9 | 0.67 | 0.65 | 0.929 |
| Alternative 4-point OGTT | 0/30/90/120 | 15*/30/90/120 | 0.42 | 57.5 | 0.69 | 0.67 | 0.945 |

WDI15–120 was recalculated using the fixed formula for each sampling schedule. Sparse curves used only retained OGTT samples, with 15\* indicating linear interpolation of the 15-min boundary. LOOCV $R^2$ reflects leave-one-out prediction of SSPG.

## Discussion

In this study, we developed and evaluated the WDI as a glucose-only OGTT metric designed to capture broad and delayed post-load glucose exposure. The principal finding is that WDI15–120 recovered substantial SSPG-related insulin-resistance information from glucose values alone.

WDI15–120 predicted SSPG more accurately in LOOCV than standard OGTT glucose summaries and established insulin-derived surrogate indices, discriminated insulin-resistant from insulin-sensitive subjects with high accuracy, and retained substantial performance when recalculated from conventional 5-point OGTT sampling. Together, these findings support WDI as a simple, physiologically interpretable glucose-only index for extracting insulin-resistance-related width and delay information from OGTT profiles.

The physiological rationale for WDI is aligned with the biology measured by SSPG. Because SSPG reflects glucose disposal capacity under comparable insulin exposure, higher SSPG values indicate greater impairment in insulin-mediated glucose disposal. During an OGTT, impaired disposal should prolong the clearance phase of the glucose excursion, causing exposure to persist across a larger fraction of the test and shifting the temporal center of exposure later. The selection of the 15–120 min window is consistent with this biology: it begins after the immediate fasting-to-post-load transition and captures the main post-load exposure and clearance period, while avoiding later recovery-phase variability that may reflect undershoot or processes less directly related to the width-delay signal. WDI captures this pattern through its relative width and *D*50_low_ components. The relative width term reflects whether glucose exposure is narrow and transient or broad and persistent after normalization to the curve’s own amplitude, while *D*50_low_ reflects whether that exposure is concentrated early or delayed later in the OGTT. The *G*_low_ scaling term further accounts for the lower envelope of the glucose profile, so that broad and delayed exposure occurring at a higher glycemic floor contributes more strongly to the index. Thus, WDI15–120 maps impaired glucose disposal onto interpretable curve features: width, delay, and floor.

WDI also differs conceptually from existing OGTT and insulin-derived surrogate indices. Standard OGTT glucose metrics, including fasting glucose, 1-h glucose, 2-h glucose, peak glucose, mean glucose, and total AUC, provide useful summaries of glycemia but do not explicitly distinguish a narrow, rapidly resolving excursion from a broad and delayed one. Total AUC captures overall exposure, but without normalization to excursion amplitude or explicit timing, it may combine biologically different curve shapes into similar values. Insulin-derived indices such as HOMA-IR, Matsuda index, and disposition index incorporate insulin information and remain important for metabolic phenotyping, but they require insulin measurements and reflect different aspects of glucose–insulin physiology. WDI is therefore not a replacement for insulin-based indices; rather, it provides a complementary glucose-only measure that extracts SSPG-relevant information from the temporal geometry of the OGTT glucose curve.

The present study uses the same dense OGTT–SSPG dataset as a previously published machine-learning analysis (Metwally, et al., 2025). That analysis showed that 16-point OGTT glucose time-series patterns contain substantial information about muscle insulin resistance, with a machine-learning model achieving an AUROC of 0.95 for insulin-resistance classification. The modeling framework used multivariable glucose-curve representations, including extracted curve features such as AUC, incremental AUC, coefficient of variation, peak glucose, time to peak, and slope-related measures, as well as reduced-representation modeling of normalized and smoothed glucose time series. In contrast, WDI was developed to reduce SSPG-related glucose-curve information to a single predefined index rather than a trained multivariable classifier. By avoiding dimensionality reduction and classifier training, WDI provides a simpler and more transparent framework for estimating SSPG-related insulin resistance from OGTT glucose profiles.

WDI has several practical and conceptual advantages. First, it is glucose-only, allowing application to OGTT datasets or clinical protocols in which insulin was not measured. This is important because glucose can be measured more easily and repeatedly than insulin, including through routine clinical testing and increasingly through continuous glucose monitoring, whereas insulin measurement requires blood sampling, laboratory assays, and greater standardization across platforms. Second, WDI is physiologically interpretable: each component corresponds to a defined curve feature, including relative exposure width, delayed exposure timing, and glycemic floor. Third, it is parameter-light and does not require kinetic modeling, curve fitting, or estimation of multiple latent variables. Finally, the sparse-sampling analysis suggests that WDI15–120 can be approximated from conventional 5-point OGTT sampling, supporting potential use beyond densely sampled research protocols. These features make WDI a practical framework for extracting dynamic insulin-resistance information from glucose profiles while preserving interpretability.

Several limitations should be noted. First, this was a secondary analysis of a relatively small cohort, and the WDI formula, window selection, and performance evaluation were derived from the same dataset; independent validation is therefore essential. Second, although SSPG provides a strong physiological reference for insulin resistance, WDI was evaluated against one reference method and may not capture all aspects of insulin sensitivity, β-cell function, or glucose tolerance. Third, the sparse-sampling analysis was performed by downsampling dense OGTT profiles rather than by prospectively collecting sparse OGTT data, so its results should be confirmed in cohorts with conventional OGTT protocols. Fourth, WDI was developed for OGTT glucose profiles; extension to other dynamic glucose settings, including mixed-meal tests or CGM-derived challenges, will require separate testing. Future studies should validate WDI in larger and more diverse cohorts, assess reproducible cutoffs, and determine whether WDI adds clinical or biological information beyond established metabolic markers.

## Conclusion

WDI provides a simple, glucose-only, and physiologically interpretable approach for extracting SSPG-related insulin-resistance information from OGTT glucose profiles. By combining relative exposure width, delayed exposure timing, and glycemic floor, WDI15–120 captured dynamic glucose features that were strongly associated with reference insulin resistance and remained informative under conventional OGTT sampling. With further validation, WDI could make routine OGTT glucose data more informative by adding a simple, interpretable measure of insulin-resistance-related curve dynamics.

## Author Contributions

R.Z. conceptualized, designed the study, analyzed the data, and wrote the paper.

## Data Availability Statement

The de-identified OGTT and SSPG data analyzed in this study are publicly available from the public repository associated with Metwally et al., Nature Biomedical Engineering 2025. Dataset: https://storage.googleapis.com/gbsc-gcp-project-ipop_public/metabolic_subphenotype_db/metabolic_subphenotypes_db.zip. Python scripts used for the analysis are available from the corresponding author upon reasonable request.

